# Protocol and statistical analysis plan for the PREOXI trial of preoxygenation with noninvasive ventilation vs oxygen mask

**DOI:** 10.1101/2023.03.23.23287539

**Authors:** Kevin W. Gibbs, Adit A. Ginde, Matthew E. Prekker, Kevin P. Seitz, Susan B. Stempek, Caleb Taylor, Sheetal Gandotra, Heath White, Daniel Resnick-Ault, Akram Khan, Amira Mohmed, Jason C. Brainard, Daniel G. Fein, Neil R. Aggarwal, Micah R. Whitson, Stephen J. Halliday, John P. Gaillard, Veronika Blinder, Brian E. Driver, Jessica A. Palakshappa, Bradley D. Lloyd, Joanne M. Wozniak, Matthew C. Exline, Derek W. Russell, Shekhar Ghamande, Cori Withers, Kinsley A. Hubel, Ari Moskowitz, Jill Bastman, Luke Andrea, Peter D. Sottile, David B. Page, Micah T. Long, Jordan Kugler Goranson, Rishi Malhotra, Brit J. Long, Steven G. Schauer, Andrew Connor, Erin Anderson, Kristin Maestas, Jillian P. Rhoads, Kelsey Womack, Brant Imhoff, David R. Janz, Stacy A. Trent, Wesley H. Self, Todd W. Rice, Matthew W. Semler, Jonathan D. Casey, the PREOXI investigators and the Pragmatic Critical Care Research Group

## Abstract

**Background:** Hypoxemia is a common and life-threatening complication during emergency tracheal intubation of critically ill adults. The administration of supplemental oxygen prior to the procedure (“preoxygenation”) decreases the risk of hypoxemia during intubation.

**Research Question:** Whether preoxygenation with noninvasive ventilation prevents hypoxemia during tracheal intubation of critically ill adults, compared to preoxygenation with oxygen mask, remains uncertain.

**Study Design and Methods:** The PRagmatic trial Examining OXygenation prior to Intubation (PREOXI) is a prospective, multicenter, non-blinded randomized comparative effectiveness trial being conducted in 7 emergency departments and 17 intensive care units across the United States. The trial compares preoxygenation with noninvasive ventilation versus oxygen mask among 1300 critically ill adults undergoing emergency tracheal intubation. Eligible patients are randomized in a 1:1 ratio to receive either noninvasive ventilation or an oxygen mask prior to induction. The primary outcome is the incidence of hypoxemia, defined as a peripheral oxygen saturation <85% between induction and 2 minutes after intubation. The secondary outcome is the lowest oxygen saturation between induction and 2 minutes after intubation. Enrollment began on 10 March 2022 and is expected to conclude in 2023.

**Interpretation:** The PREOXI trial will provide important data on the effectiveness of noninvasive ventilation and oxygen mask preoxygenation for the prevention of hypoxemia during emergency tracheal intubation. Specifying the protocol and statistical analysis plan prior to the conclusion of enrollment increases the rigor, reproducibility, and interpretability of the trial.

**Clinical trial registration number:** NCT05267652

**HIGHLIGHTS:** • Hypoxemia is common during emergency tracheal intubation

• Supplemental oxygen prior to intubation (preoxygenation) reduces risk of hypoxemia

• The PREOXI trial compares noninvasive ventilation vs oxygen mask preoxygenation

• This protocol describes the design, methods, and planned analyses

• PREOXI is the largest trial of preoxygenation for emergency intubation to date

## INTRODUCTION

Life-threatening hypoxemia occurs in 10-20% of emergency tracheal intubations in the Emergency Department (ED) and Intensive Care Unit (ICU).^1, 2^ Hypoxemia during intubation is associated with an increased risk of cardiac arrest and death.^3, 4^ Identifying interventions to prevent hypoxemia during emergency tracheal intubation is a high priority for clinicians and researchers.^5, 6^ Because patients are typically apneic between induction of anesthesia and intubation but continue to consume oxygen, the oxygen content in the lungs at the time of induction is a primary determinant of whether the patient will experience hypoxemia. Preoxygenation, the administration of supplemental oxygen prior to induction of anesthesia, increases the oxygen content in the lung at induction and decreases the risk of hypoxemia.^7, 8^ In current clinical practice, preoxygenation for emergency tracheal intubation of critically ill adults is most commonly administered using either an oxygen mask or noninvasive ventilation.^1^

Preoxygenation with an oxygen mask is typically performed using either a non-rebreather mask or a bag-mask device. A non-rebreather mask is a loose-fitting mask with an oxygen reservoir connected to an oxygen source. A bag-mask device is a mask capable of forming a tight seal over the mouth when held in place by a clinician and can be used to provide supplemental oxygenation alone, or both supplemental oxygen and manual ventilation.^9^ Both types of oxygen mask can deliver up to 100% oxygen, are simple to set up, and have low potential for gastric insufflation. However, oxygen masks may deliver oxygen less effectively in critically ill patients when tachypnea, high minute ventilation, and poor mask seal allow the entrainment of ambient air with resulting alveolar oxygen concentrations as low as 50%.^7, 10^

Preoxygenation is also routinely administered via noninvasive ventilation, in which a tight-fitting mask is connected to a machine capable of providing both 100% oxygen and positive pressure ventilation. Compared to an oxygen mask, noninvasive ventilation may reduce air entrainment by delivering a higher inspiratory flow rate of oxygen and by minimizing leaks. Additionally, noninvasive ventilation increases the mean airway pressure and recruits atelectatic lung, potentially reducing shunting. Compared with an oxygen mask, noninvasive ventilation may take longer to initiate and may potentially increase the risk of gastric insufflation and aspiration.

Two small, randomized trials have compared these two approaches to preoxygenation. The first trial found that, among 53 ICU patients undergoing tracheal intubation in two hospitals in France, noninvasive ventilation increased the lowest oxygen saturation compared to an oxygen mask (mean lowest oxygen saturation 93% vs. 81%, respectively, P<0.001) with no difference between groups in the incidence of aspiration (6% vs. 8%).^11^ Among 201 ICU patients in 6 hospitals in France, the second trial found no difference in the severity of illness in the 7 days after intubation and an incidence of hypoxemia during intubation of 18.4% in the noninvasive ventilation group versus 27.7% in the oxygen mask group (P = 0.10).^12^ Thus, whether noninvasive ventilation for preoxygenation in critically ill adults undergoing emergency tracheal intubation decreases the incidence of hypoxemia compared to an oxygen mask remains unknown. Therefore, we designed the PRagmatic trial Examining OXygenation prior to Intubation (PREOXI) to test the hypothesis that, among critically ill adults undergoing emergency tracheal intubation in the ED and ICU, preoxygenation with noninvasive ventilation will decrease the incidence of hypoxemia compared to preoxygenation with an oxygen mask.

## METHODS AND ANALYSIS

This manuscript was written in accordance with Standard Protocol Items: Recommendations for Interventional Trials (SPIRIT) guidelines (see Table 1 and online supplement file 1, section 1). ^13^

**Table 1:** Schedule of Enrollment, Interventions, and Assessments in PREOXI.

|  |  |  |  |  |  |  |  |
| --- | --- | --- | --- | --- | --- | --- | --- |
|  | Eligibility Screen | Randomization and Allocation | Peri-Procedural |  |  |  | Final Outcome Assessment |
| Timepoint | Decision to perform tracheal intubation | Before tracheal intubation | Induction | Tracheal intubation | 0-2 min after tracheal intubation | 0-24 hours after tracheal intubation | Discharge or 28 days after enrollment |
| Enrollment: |  | X |  |  |  |  |  |
| Eligibility screen | X |  |  |  |  |  |  |
| Allocation |  | X |  |  |  |  |  |
| Interventions: |  |  |  |  |  |  |  |
| Preoxygenation with NIPPV |  | X | X |  |  |  |  |
| Preoxygenation with facemask oxygen |  | X | X |  |  |  |  |
| Screening for contraindications | X | X |  |  |  |  |  |
| Assessments: |  |  |  |  |  |  |  |
| Baseline variables | X | X |  |  |  |  |  |
| Peri-procedural variables |  | X | X | X | X |  |  |
| Adverse events |  | X | X | X | X | X | X |
| Clinical outcomes |  |  |  |  | X | X | X |

### Funding

Funding for this trial was provided by the Department of Defense, Defense Health Agency, J9 Office, RESTORAL program. The funder has no role in study design or conduct, data collection or analysis.

### Patient and public involvement

Materials used to communicate about the study with patients and families were developed with input from the Vanderbilt Community Engaged Research Core, which includes input from patients and community members. Study authors will disseminate the results of this study online and via social media in forms suitable for public understanding.

### Study Design

PREOXI is a pragmatic, multicenter, non-blinded, parallel-group, randomized trial comparing preoxygenation with noninvasive ventilation to preoxygenation with an oxygen mask among critically ill adults undergoing emergency tracheal intubation in the ED and ICU. The primary outcome is the incidence of hypoxemia, defined as a peripheral oxygen saturation < 85% between induction of anesthesia and two minutes after intubation. The trial is conducted by the Pragmatic Critical Care Research Group (www.pragmaticcriticalcare.org). An independent data and safety monitoring board (DSMB) is monitoring the progress and safety of the trial. The trial was registered prior to initiation of enrollment (ClinicalTrials.gov identifier: NCT05267652).

### Study Population

Patients located in a participating ED or ICU who are undergoing tracheal intubation using a laryngoscope and sedation are eligible. Patients are excluded if they are known to be less than 18 years old, are known to be pregnant or a prisoner, require positive pressure ventilation for apnea or hypopnea, or have an immediate need for tracheal intubation that precludes performance of study procedures, or if the clinician performing the procedure (referred to as the “operator”) determines that preoxygenation with noninvasive ventilaton or an oxygen mask is either required or contraindicated. Complete lists of inclusion and exclusion criteria are provided in Table 2.

**Table 2:** Inclusion/Exclusion Criteria.

|  |  |
| --- | --- |
| **Inclusion Criteria** |  |
|  | Patient is located in a participating unit. |
|  | Planned procedure is tracheal intubation using a laryngoscope and sedation. |
|  | Planned operator is a clinician expected to routinely perform tracheal intubation in the participating unit. |
| **Exclusion Criteria** |  |
|  | Patient is receiving positive pressure ventilation by a mechanical ventilator, bag-mask device, or laryngeal mask airway. |
|  | Patient is known to be less than 18 years old. |
|  | Patient is known to be pregnant. |
|  | Patient is known to be a prisoner. |
|  | Immediate need for tracheal intubation precludes safe performance of study procedures. |
|  | Patient is apneic, hypopneic, or has another condition requiring positive pressure ventilation between enrollment and induction. |
|  | Operator has determined that preoxygenation with noninvasive ventilation or preoxygenation with an oxygen mask is required or contraindicated for optimal care of the patient. |

### Randomization and treatment allocation

Patients are randomized in a 1:1 ratio to undergo preoxygenation with noninvasive ventilaton vs oxygen mask in permuted blocks of variable size, stratified by study site. Study-group assignments are generated using a computerized randomization sequence, placed in sequentially numbered opaque envelopes, and distributed to enrolling sites. Before opening the envelope, the operator determines that the patient meets eligibility criteria. Study group assignment remains concealed to study personnel and treating clinicians until after the decision has been made to enroll the patient and the envelope is opened. Patients are considered to be enrolled once the operator opens the opaque trial envelope to reveal study group assignment. After randomization, patients, treating clinicians, and study personnel are not blinded to study group assignment due to the nature of the study intervention.

## STUDY INTERVENTIONS

### Training

Before beginning enrollment at each site, study investigators provide training on study procedures including instructional videos with consensus best practice recommendations for preoxygenation with noninvasive ventilation and preoxygenation with an oxygen mask. Observers receive in-person training on the collection of data during the intubation procedure using example data collection sheets. Descriptions of the training videos and website links can be found in the supplementary appendix.

### Noninvasive ventilation group

For patients assigned to the noninvasive ventilation group, operators are instructed to administer noninvasive ventilation via a tight-fitting mask covering the nose and mouth connected to either a conventional mechanical ventilator or a dedicated noninvasive ventilator (i.e., BiPAP machine) from the initiation of preoxygenation until the initiation of laryngoscopy. The trial protocol does not dictate brand or type of ventilator, ventilator settings, or duration of preoxygenation. Operators receive the following best practice recommendations for the administration of preoxygenation using noninvasive ventilation:

1. Preoxygenate ≥ 3 minutes (if feasible)
2. Continue noninvasive ventilation until initiation of laryngoscopy
3. Fraction of Inspired Oxygen (FiO2) of 100%
4. Expiratory pressure ≥ 5 cm of water
5. Inspiratory pressure ≥ 10 cm of water
6. Respiratory rate of ≥ 10

### Oxygen mask group

For patients assigned to the oxygen mask group, clinicians are instructed to administer supplemental oxygen via a non-rebreather mask or bag-mask device without manual ventilation from the initiation of preoxygenation until the induction of anesthesia. The operator determines whether to use a non-rebreather mask or a bag-mask device without manual ventilation. The trial protocol does not dictate the brand or type of non-rebreather or bag-mask device or duration of preoxygenation. Between induction of anesthesia and initiation of laryngoscopy, the operator determines whether to provide oxygen with a non-rebreather mask, oxygen with a bag-mask device without manual ventilation, or oxygen with a bag-mask device with manual ventilation.^2^ Operators receive the following best practice recommendations for the administration of preoxygenation using an oxygen mask:

1. Preoxygenate ≥ 3 minutes (if feasible)
2. Maximal oxygen flow rate possible (≥ 15 liters per minute)
3. Continue oxygenation from induction to laryngoscopy

### Cointerventions

Study group assignment determines only the initial method of preoxygenation. Treating clinicians determine all other aspects of the intubation procedure including: [1] the co-administration of supplemental oxygen by nasal cannula (either standard nasal cannula, large bore nasal cannula, or heated high flow nasal cannula) before induction, between induction an initiation of laryngoscopy, and between initiation of laryngoscopy and intubation of the trachea; [2] choice of induction medication and timing of administration; [3] use of neuromuscular blockade; [4] choice of laryngoscope; [5] use of additional airway management equipment and adjuncts; and [6] post-intubation ventilator settings.

### Data Collection

An observer not directly involved with the intubation procedure collects data for key periprocedural outcomes, including oxygen saturation at induction and the lowest value for oxygen saturation between induction and two minutes after successful intubation. Observers may be clinical personnel on the enrolling unit (e.g., physicians or nurses) or research personnel. Immediately after the intubation procedure, the operator completes a paper data collection form to record the device used for preoxygenation, the duration of preoxygenation, the devices used for oxygenation and ventilation between induction and laryngoscopy, and complications of intubation.^14^ Study personnel at each site review the medical record to collect data on baseline characteristics, pre-and post-laryngoscopy management, and clinical outcomes. A complete list of baseline, peri-procedural, and in-hospital variables are provided in Supplemental Table 1.

Data on pneumothorax and new pulmonary infiltrates are collected by study staff from clinical radiology reports using a structured case report form. The clinical radiologist who interprets the chest imaging is unaware of study group assignment. A lung infiltrate is considered to be present if the clinical radiologist identifies on the chest imaging the presence of air bronchograms, centrilobular nodules, consolidation, ground-glass opacity, infiltrate, opacity, parenchymal opacification, pneumonia, pneumonitis pulmonary edema, or a tree-in-bud pattern. A pneumothorax or lung infiltrate present on chest imaging in the 24 hours after intubation will be assumed to be new if it was not present on chest imaging in the 24 hours prior to intubation. If no chest imaging is available in the 24 hours prior to intubation, any pneumothorax or lung infiltrate on chest imaging will be assumed to be new.

### Primary Outcome

The primary outcome is the incidence of hypoxemia, defined as a peripheral oxygen saturation <85% during the interval between induction and 2 minutes after intubation.

We selected hypoxemia (as a binary variable) rather than lowest oxygen saturation (a continuous variable) as the primary outcome for the trial for several reasons. First, experiencing hypoxemia in the range associated with an increased risk of adverse clinical outcomes (e.g., cardiac arrest) may be more clinically relevant than changes in oxygen saturation within the normal range. For example a change in oxygen saturation of 5 percentage points from 87% to 82% may be more closely associated with adverse outcomes than a change in oxygen saturation of 10 percentage points from 100% to 90%. Second, values for oxygen saturation are “right-censored” because oxygen saturation reaches 100% with a partial pressure of oxygen in arterial blood (PaO_2_) of approximately 100 mmHg but patients may have a higher PaO_2_ following preoxygenation. The approach of analyzing hypoxemia as a binary variable rather than lowest oxygen saturation has been used by many prior trials and endorsed by airway experts.^15–20^

We selected an oxygen saturation of <85% as the threshold for the primary outcome based on several physiologic and procedural factors. First, an oxygen saturation of 85% corresponds with the inflection point on the oxyhemoglobin dissociation curve, at which further decrements in arterial oxygen concentrations result in rapid and critical desaturation.^9^ Second, an oxygen saturation <85% has been associated with an increased risk of cardiac arrest during tracheal intubation^21^ and may be associated with increased mortality.^5^

### Secondary Outcome

The sole secondary outcome is the lowest oxygen saturation during the interval between induction and 2 minutes after tracheal intubation.

### Additional Outcomes

Table 3 reports the safety outcomes, exploratory outcomes, and clinical outcomes.

**Table 3:** Study outcomes.

|  |  |
| --- | --- |
| **Primary Outcome** | Incidence of hypoxemia, defined as a peripheral oxygen saturation <85% during the interval between induction and 2 minutes after tracheal intubation. |
| **Secondary Outcome** | Lowest oxygen saturation during the interval between induction and 2 minutes after tracheal intubation. |
| **Safety Outcomes** | Incidence of operator-reported aspiration. |
|  | Fraction of inspired oxygen at 24 hours after induction. |
|  | Oxygen saturation at 24 hours after induction. |
|  | Incidence of pneumothorax, defined as radiology report of new pneumothorax on chest imaging in the 24 hours after induction. If no chest imaging available pre-induction, any pneumothorax in the 24 hours after induction will be assumed to be new. |
|  | New infiltrate, defined as radiology report of new infiltrate on chest imaging in the 24 hours after intubation. If no chest imaging available pre-induction, any infiltrate in the 24 hours after induction will be assumed to be new. |
| **Exploratory Outcomes** | Incidence of severe hypoxemia, defined as the lowest oxygen saturation of <80% between induction and 2 minutes after tracheal intubation. |
|  | Incidence of very severe hypoxemia, defined as the lowest oxygen saturation of <70% between induction and 2 minutes after tracheal intubation. |
|  | Oxygen saturation at induction. |
|  | Systolic blood pressure at induction. |
|  | Duration from induction to successful intubation (duration of the intubation procedure). |
|  | Cormack-Lehane grade of glottic view on first attempt. |
|  | Number of laryngoscopy attempts. |
|  | Number of attempts at passing a bougie. |
|  | Number of attempts at passing an endotracheal tube. |
|  | Incidence of cardiovascular collapse, defined as a composite of one or more of the following between induction and 2 minutes after intubation: Systolic blood pressure <65 mmHg, new or increased vasopressor, cardiac arrest not resulting in death within 1 hour of induction, cardiac arrest resulting in death within 1 hour of induction. |
|  | 28 day in-hospital mortality. |
|  | Ventilator-free days to 28 days. |
|  | ICU-free days to 28 days. |

### Sample Size Estimation

The minimum clinically important difference in the incidence of hypoxemia that would be required to justify routine preoxygenation with noninvasive ventilation rather than preoxygenation with an oxygen mask during the emergency tracheal intubation of critically ill adults is uncertain. The current trial is designed to detect a 6% absolute difference between groups in the incidence of hypoxemia, a difference that is similar to or smaller than the difference considered to be clinically meaningful in the design of prior trials of oxygenation strategies during tracheal intubation.^2, 24^ Assuming an incidence of hypoxemia of 17% in the oxygen mask group based on data from two recently completed trials by this network in similar ED and ICU settings, detecting a 6% absolute decrease in the incidence of hypoxemia with 85% power at a two-sided alpha level of 0.05 would require enrollment of 1,264 patients (632 per group).^25, 26^ Anticipating missing data for up to 3% of patients, we will plan to enroll a maximum of 1,300 total patients (650 per group). This sample size calculation was performed in PS version (Nashville, Tennessee).

### Data and Safety Monitoring Board and Interim Analysis

A data and safety monitoring board (DSMB) consisting of members with expertise in bioethics, emergency medicine, pulmonary and critical care medicine, anesthesiology, biostatistics, and clinical trial methodology has overseen the design of the trial and is monitoring its conduct. The DSMB will review a single interim analysis, prepared by the study biostatistician at the anticipated halfway point of the trial after enrolment of 650 patients. The pre-specified stopping boundary for efficacy is a P value < 0.001 using a Chi-square test for the difference between groups in the primary outcome. This conservative Haybittle–Peto boundary will allow the final analysis to be performed using an unchanged level of significance (two-sided P value < 0.05).

The DSMB has the authority to recommend that the trial stop at any point, request additional data, request additional interim analyses, or request modifications to the study protocol.

### Statistical Analysis Principles

Analyses will be conducted following reproducible research principles using R (R Foundation for Statistical Computing, Vienna, Austria). Categorical variables will be presented as number and percentage and compared between groups using a Chi-square text. Continuous variables will be presented as mean ± SD or median and IQR and compared between groups using a Wilcoxon Rank Sum test. We will also present absolute between-group differences with associated 95% confidence intervals. A two-sided P-value of < 0.05 will define a statistically significant between-group difference in the primary outcome. With a single primary outcome, no adjustment for multiplicity will be made. For secondary, safety, and exploratory analyses, emphasis will be placed on the magnitude of differences between groups with 95% confidence intervals rather than statistical significance.

### Main Analysis of the Primary Outcome

The main analysis will be an unadjusted, intention-to-treat comparison of the primary outcome of hypoxemia between patients randomized to the noninvasive ventilation group versus patients randomized to the oxygen mask group, using a chi-square test. The absolute difference in proportions, associated 95% confidence interval, and a P value for the comparison will be presented. The primary analysis will be conducted among patients for whom the primary outcome is available without imputation of missing data.

### Additional Analyses of the Primary Outcome

#### Multivariable analysis

To account for relevant baseline covariates, we will fit a generalized linear mixed effects model using a logit link function with the primary outcome as the dependent variable, study site as a random effect, and fixed effects of study group and the following prespecified baseline covariates: age, sex, race and ethnicity, body mass index (BMI), location at enrollment (ED or ICU), highest fraction of inspired oxygen in the hour prior to initiation of preoxygenation, Acute Physiology and Chronic Health Evaluation (APACHE) II score^27^, and indication for intubation (hypoxemic respiratory failure: Yes vs No). All continuous variables will be modelled assuming a non-linear relationship to the outcome using restricted cubic splines with between 3 and 5 knots.

#### Effect modification

We will examine whether prespecified baseline variables modify the effect of study group assignment (noninvasive ventilation vs oxygen mask) on the primary outcome using a formal test of statistical interaction in a generalized linear mixed-effects model with the primary outcome as the dependent variable, study site as a random effect and fixed effects of study group, the prespecified proposed effect modifier and the interaction between the two. For categorical variables, we will present the OR and 95% CIs within each prespecified subgroup. Continuous variables will not be dichotomized for analysis of effect modification but may be dichotomized for data presentation. In accordance with the Instrument for assessing the Credibility of effect Modification Analyses (ICEMAN) recommendations, we have prespecified the following baseline variables as potential effect modifiers and hypothesized the direction of effect modification for each:

1. <u>Patient location (ED vs ICU)</u>. We hypothesize that patient location will not modify the effect of study group assignment on the primary outcome.
2. Body Mass Index (kg/m^2^). We hypothesize that Body Mass Index (BMI) will modify the effect of study group assignment on the primary outcome, with a greater decrease in the incidence of hypoxemia with preoxygenation within the noninvasive ventilation group compared to the oxygen mask group among patients with higher BMIs, as compared to patients with lower BMIs. This hypothesis of effect modification is supported by evidence from multiple prior studies that patients with obesity are more likely to have early airway closure and atelectasis-dependent shunting that is likely to improve with positive pressure ventilation. ^28, 29^
3. <u>Fraction of inspired oxygen in the hour prior to intubation</u>. We hypothesize that the fraction of inspired oxygen received in the hour prior to intubation will modify the effect of study group assignment on the primary outcome, with a greater decrease in the incidence of hypoxemia in the noninvasive ventilation group compared to the oxygen mask group among patients with higher fractions of inspired oxygen in the hour prior to intubation, compared to patients with lower fractions of inspired oxygen. This hypothesis of effect modification is supported by evidence from multiple prior studies that patients requiring higher fractions of inspired oxygen have more atelectasis-dependent shunting that is likely to improve with positive pressure ventilation.^5, 11, 30^
4. <u>APACHE II score</u>. We hypothesize that APACHE II score will not modify the effect of study group assignment on the primary outcome.
5. <u>Hypoxemic respiratory failure as the indication for intubation (Yes vs No)</u>. We hypothesize that hypoxemic respiratory failure as the indication for intubation will modify the effect of study group assignment on the primary outcome, with a greater decrease in the incidence of hypoxemia in the noninvasive ventilation group compared to the oxygen mask group among patients with hypoxemic respiratory failure, compared to patients without. This hypothesis of effect modification is supported by evidence from two prior randomized trials suggesting a potential benefit for noninvasive ventilation among patients with acute hypoxemic respiratory failure.^11, 12^

### Analysis of the Secondary Outcome

We will perform an unadjusted, intention-to-treat comparison of patients randomized to the noninvasive ventilation group versus the oxygen mask group with regard to the secondary outcome of lowest oxygen saturation between induction and 2 minutes after intubation using the Wilcoxon rank sum test.

### Analyses of Additional Outcomes

We will conduct unadjusted, intention-to-treat analyses comparing patients randomized to the noninvasive ventilation group versus the oxygen mask group with regard to all pre-specified safety, clinical, and exploratory outcomes. Continuous outcomes will be compared with the Wilcoxon rank sum test and categorical variables with the chi-square test. Between-group differences in continuous and categorical variables and the associated 95% confidence intervals will be presented.

### Handling of missing data

We anticipate that no patients will be lost to follow up before assessment of the primary outcome. In some cases, the oxygen saturation between induction and 2 minutes after intubation will be unmeasurable (e.g., poor plethysmography of pulse oximetry, shock, cardiac arrest, peripheral arterial disease, or other reasons) or unavailable. We anticipate that data will be missing in less than 3% of cases based on the rates of missing data in prior trials in similar settings.^25, 26^ Missing data will not be imputed for the primary outcome or for any of the secondary or exploratory outcomes. In adjusted analyses, missing data for baseline covariates will be imputed using multiple imputations.

### Trial status

PREOXI is a pragmatic, multi-center, non-blinded randomized clinical trial comparing preoxygenation with noninvasive ventilation to preoxygenation with an oxygen mask during the tracheal intubation of critically ill adults. Enrollment began on 10 March 2022 and is expected to conclude in 2023.

## ETHICS AND DISSEMINATION

### Waiver of Informed Consent

Critically ill patients undergoing tracheal intubation in the ED or ICU are at significant risk for morbidity and mortality from their underlying illness. Most patients undergoing tracheal intubation in routine clinical care receive preoxygenation with either noninvasive ventilation or an oxygen mask. Any benefits or risks of these two approaches are experienced by patients undergoing tracheal intubation in clinical care, outside the context of research. As a requirement for enrollment in PREOXI, the patient’s treating clinician must determine that either preoxygenation with noninvasive ventilation or preoxygenation with an oxygen mask would be a safe and reasonable approach for the patient (otherwise the patient is excluded). Therefore, making the decision between the two approaches randomly in the context of a pragmatic trial, rather than by a clinician who thinks either approach is safe and reasonable for the patient, is expected to pose no more than minimal additional risk.

Obtaining informed consent from potential study participants or their legally authorized representatives would be impracticable. The majority of critically ill patients undergoing emergency tracheal intubation lack decisional capacity due to their critical illness and surrogate decision makers are frequently unavailable. Further, emergency tracheal intubation is a time-sensitive procedure with only minutes between the decision to intubate and the completion of the procedure. Meaningful informed consent could not be executed in this brief window. Attempting to obtain informed consent could lead to potentially deleterious delays in intubation which would increase the risk of hypoxemia, hypotension, and cardiac arrest.

Because the study involves minimal incremental risk, the study would not adversely affect the welfare or privacy rights of the participant, and obtaining informed consent would be impracticable, a waiver of informed consent was requested from and approved by the single institutional review board at Vanderbilt University Medical Center (reference number VUMC IRB# 211271). Conduct of this trial with waiver of informed consent is consistent with previous randomized trials comparing alternative approaches to tracheal intubation in widespread clinical use.^2, 24–26, 31, 32^ This approach was secondarily approved by the US Department of Defense Health Agency Human Research Protection Office). IRBs at participating sites reviewed the protocol, addressed any local contextual factors with the site principal investigator, and ceded responsibility for ethics approval and study oversight to the single IRB.

### Information for Patients and Families

Information regarding the study is made available to patients and families following intubation using a patient and family information sheet. The patient and family information sheet contains information on the purpose of the PREOXI trial, study procedures, risks and discomforts, benefits, use of protected health information, confidentiality, and investigator contact information. The Defense Health Agency Human Research Protection Office determined that this procedure meets the requirement of 32 CFR 219 and DODI 3216.02_AFI40-402. At centers with a significant population of non-English speaking patients, the patient and family information sheet has been translated into Spanish and Somali languages.

### Protocol Changes

In accordance with SPIRIT guidelines, changes to the study protocol will be documented on clinicaltrials.gov (see online supplementary file) and submitted to the sIRB for approval.

### Data Handling

Privacy protocols and data handling are reported in the online supplement.

### Dissemination Plan

We will submit the trial results to a peer-reviewed journal for publication. Trial results will also be presented at scientific conferences and disseminated online and via social media in forms suitable for public understanding.

### Conclusion

The PREOXI trial will provide important data on the effectiveness of common preoxygenation strategies for the prevention of hypoxemia during emergency tracheal intubation with a goal of improving outcomes for critically ill adults. To aid in the transparency and interpretation of trial results, this protocol and statistical analysis plan for the PREOXI trial has been finalized prior to the conclusion of patient enrollment.

## Author contributions

Study concept and design: KWG, AAG, MWS, JDC; Acquisition of data: all authors; Drafting of the manuscript KWG, AAG, MWS, JDC; Critical revision of the manuscript for important intellectual content: all authors; Study supervision: KWG, AAG, MWS, JDC.

## Supporting information

PREOXI protocol supplement

## Data Availability

All data produced in the present study are available upon reasonable request to the authors

## REFERENCES

1. Russotto V, Myatra SN, Laffey JG, et al. Intubation Practices and Adverse Peri-intubation Events in Critically Ill Patients From 29 Countries. JAMA. Mar 23 2021;325(12):1164–1172. doi:10.1001/jama.2021.1727

2. Casey JD, Janz DR, Russell DW, et al. Bag-Mask Ventilation during Tracheal Intubation of Critically Ill Adults. N Engl J Med. Feb 28 2019;380(9):811–821. doi:10.1056/NEJMoa1812405

3. Davis DP, Dunford JV, Poste JC, et al. The impact of hypoxia and hyperventilation on outcome after paramedic rapid sequence intubation of severely head-injured patients. J Trauma. Jul 2004;57(1):1–8; discussion 8-10. doi:10.1097/01.ta.0000135503.71684.c8

4. April MD, Arana A, Reynolds JC, et al. Peri-intubation cardiac arrest in the Emergency Department: A National Emergency Airway Registry (NEAR) study. Resuscitation. May 2021;162:403–411. doi:10.1016/j.resuscitation.2021.02.039

5. Smischney NJ, Khanna AK, Brauer E, et al. Risk Factors for and Outcomes Associated With Peri-Intubation Hypoxemia: A Multicenter Prospective Cohort Study. J Intensive Care Med. Dec 2021;36(12):1466–1474. doi:10.1177/0885066620962445

6. Meier NM, Gibbs KW. Evolving Tracheal Intubation Practice Patterns in the Pandemic Era. Chest. Dec 2021;160(6):1993–1994. doi:10.1016/j.chest.2021.07.002

7. Mosier JM, Hypes CD, Sakles JC. Understanding preoxygenation and apneic oxygenation during intubation in the critically ill. Intensive Care Med. Feb 2017;43(2):226–228. doi:10.1007/s00134-016- 4426-0

8. Berthoud M, Read DH, Norman J. Pre-oxygenation--how long? Anaesthesia. Feb 1983;38(2):96–102. doi:10.1111/j.1365-2044.1983.tb13925.x

9. Weingart SD. Preoxygenation, reoxygenation, and delayed sequence intubation in the emergency department. J Emerg Med. Jun 2011;40(6):661–7. doi:10.1016/j.jemermed.2010.02.014

10. Driver BE, Prekker ME, Kornas RL, Cales EK, Reardon RF. Flush Rate Oxygen for Emergency Airway Preoxygenation. Ann Emerg Med. Jan 2017;69(1):1–6. doi:10.1016/j.annemergmed.2016.06.018

11. Baillard C, Fosse JP, Sebbane M, et al. Noninvasive ventilation improves preoxygenation before intubation of hypoxic patients. Am J Respir Crit Care Med. Jul 15 2006;174(2):171–7. doi:10.1164/rccm.200509-1507OC

12. Baillard C, Prat G, Jung B, et al. Effect of preoxygenation using non-invasive ventilation before intubation on subsequent organ failures in hypoxaemic patients: a randomised clinical trial. Br J Anaesth. Feb 2018;120(2):361–367. doi:10.1016/j.bja.2017.11.067

13. Chan AW, Tetzlaff JM, Gotzsche PC, et al. SPIRIT 2013 explanation and elaboration: guidance for protocols of clinical trials. BMJ. Jan 8 2013;346:e7586. doi:10.1136/bmj.e7586

14. Cormack RS, Lehane J. Difficult tracheal intubation in obstetrics. Anaesthesia. Nov 1984;39(11):1105–11.

15. Writing C, Steering Committee for the RCG, Algera AG, et al. Effect of a Lower vs Higher Positive End-Expiratory Pressure Strategy on Ventilator-Free Days in ICU Patients Without ARDS: A Randomized Clinical Trial. JAMA. Dec 22 2020;324(24):2509–2520. doi:10.1001/jama.2020.23517

16. Franklin D, Babl FE, Schlapbach LJ, et al. A Randomized Trial of High-Flow Oxygen Therapy in Infants with Bronchiolitis. N Engl J Med. Mar 22 2018;378(12):1121–1131. doi:10.1056/NEJMoa1714855

17. Driver BE, Semler MW, Self WH, et al. Effect of Use of a Bougie vs Endotracheal Tube With Stylet on Successful Intubation on the First Attempt Among Critically Ill Patients Undergoing Tracheal Intubation: A Randomized Clinical Trial. JAMA. Dec 28 2021;326(24):2488–2497. doi:10.1001/jama.2021.22002

18. De Jong A, Rolle A, Molinari N, et al. Cardiac Arrest and Mortality Related to Intubation Procedure in Critically Ill Adult Patients: A Multicenter Cohort Study. Crit Care Med. Apr 2018;46(4):532–539. doi:10.1097/CCM.0000000000002925

19. Zhang C, Xu F, Li W, et al. Driving Pressure-Guided Individualized Positive End-Expiratory Pressure in Abdominal Surgery: A Randomized Controlled Trial. Anesth Analg. Nov 1 2021;133(5):1197–1205. doi:10.1213/ANE.0000000000005575

20. Kritek PA, Luks AM. Preventing Dogma from Driving Practice. N Engl J Med. Feb 28 2019;380(9):870–871. doi:10.1056/NEJMe1900708

21. Mort TC. The incidence and risk factors for cardiac arrest during emergency tracheal intubation: a justification for incorporating the ASA Guidelines in the remote location. J Clin Anesth. Nov 2004;16(7):508–16. doi:10.1016/j.jclinane.2004.01.007

22. Higgs A, McGrath BA, Goddard C, et al. Guidelines for the management of tracheal intubation in critically ill adults. Br J Anaesth. Feb 2018;120(2):323–352. doi:10.1016/j.bja.2017.10.021

23. Brown CA, 3rd, Mosier JM, Carlson JN, Gibbs MA. Pragmatic recommendations for intubating critically ill patients with suspected COVID-19. J Am Coll Emerg Physicians Open. Apr 2020;1(2):80–84. doi:10.1002/emp2.12063

24. Semler MW, Janz DR, Lentz RJ, et al. Randomized Trial of Apneic Oxygenation during Endotracheal Intubation of the Critically Ill. Am J Respir Crit Care Med. Feb 1 2016;193(3):273–80. doi:10.1164/rccm.201507-1294OC

25. Driver BE, Prekker ME, Casey JD. Use of a Bougie vs Endotracheal Tube With Stylet and Successful Intubation on the First Attempt Among Critically Ill Patients Undergoing Tracheal Intubation-Reply. JAMA. Apr 19 2022;327(15):1503–1504. doi:10.1001/jama.2022.2716

26. Russell DW, Casey JD, Gibbs KW, et al. Effect of Fluid Bolus Administration on Cardiovascular Collapse Among Critically Ill Patients Undergoing Tracheal Intubation: A Randomized Clinical Trial. JAMA. Jul 19 2022;328(3):270–279. doi:10.1001/jama.2022.9792

27. Knaus WA, Draper EA, Wagner DP, Zimmerman JE. APACHE II: a severity of disease classification system. Crit Care Med. Oct 1985;13(10):818–29.

28. Delay JM, Sebbane M, Jung B, et al. The effectiveness of noninvasive positive pressure ventilation to enhance preoxygenation in morbidly obese patients: a randomized controlled study. Anesth Analg. Nov 2008;107(5):1707–13. doi:10.1213/ane.0b013e318183909b

29. Grieco DL, Anzellotti GM, Russo A, et al. Airway Closure during Surgical Pneumoperitoneum in Obese Patients. Anesthesiology. Jul 2019;131(1):58–73. doi:10.1097/ALN.0000000000002662

30. Coussa M, Proietti S, Schnyder P, et al. Prevention of atelectasis formation during the induction of general anesthesia in morbidly obese patients. Anesth Analg. May 2004;98(5):1491–5, table of contents. doi:10.1213/01.ane.0000111743.61132.99

31. Janz DR, Casey JD, Semler MW, et al. Effect of a fluid bolus on cardiovascular collapse among critically ill adults undergoing tracheal intubation (PrePARE): a randomised controlled trial. Lancet Respir Med. Dec 2019;7(12):1039–1047. doi:10.1016/S2213-2600(19)30246-2

32. Janz DR, Semler MW, Lentz RJ, et al. Randomized Trial of Video Laryngoscopy for Endotracheal Intubation of Critically Ill Adults. Crit Care Med. Nov 2016;44(11):1980–1987. doi:10.1097/CCM.0000000000001841

