## Supplementary material for "Protocol and statistical analysis plan for the PREOXI trial of preoxygenation with noninvasive ventilation vs oxygen mask": PREOXI protocol supplement

Table of Contents

1. SPIRIT 2013 Checklist
2. List of PREOXI Investigators
3. Definition of ICU-Free Days (ICU-FDs)
4. Definition of Ventilator-Free Days (VFDs)
5. Safety Monitoring and Adverse Events

5.1.	Adverse Event Definitions

5.2.	Monitoring for Adverse Events

5.3	Recording and Reporting Adverse Events

5.4	Clinical Outcomes that may be Exempt from Adverse Event Recording and Reporting

5.5	Unanticipated Problems involving Risks to Subjects or Others

6.	Patient Privacy and Data Storage

7. Plan for communication of Protocol Changes

8.	Training materials

9. Supplementary Table

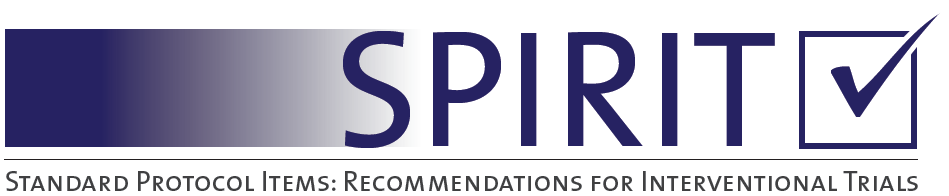

SPIRIT 2013 Checklist: Recommended items to address in a clinical trial protocol and related documents*

| Section/item | Item  No | Description | Addressed on page number |
| --- | --- | --- | --- |
| **Administrative information** | | |  |
| Title | 1 | Descriptive title identifying the study design, population, interventions, and, if applicable, trial acronym | __1__ |
| Trial registration | 2a | Trial identifier and registry name. If not yet registered, name of intended registry | __6__ |
|  | 2b | All items from the World Health Organization Trial Registration Data Set | __1-6__ |
| Protocol version | 3 | Date and version identifier | __NA__ |
| Funding | 4 | Sources and types of financial, material, and other support | __5__ |
| Roles and responsibilities | 5a | Names, affiliations, and roles of protocol contributors | __1-3__ |
|  | 5b | Name and contact information for the trial sponsor | __1-5__ |
|  | 5c | Role of study sponsor and funders, if any, in study design; collection, management, analysis, and interpretation of data; writing of the report; and the decision to submit the report for publication, including whether they will have ultimate authority over any of these activities | __1- 5__ |
|  | 5d | Composition, roles, and responsibilities of the coordinating centre, steering committee, endpoint adjudication committee, data management team, and other individuals or groups overseeing the trial, if applicable (see Item 21a for data monitoring committee) | __1-5, *__ |
| Introduction |  |  |  |
| Background and rationale | 6a | Description of research question and justification for undertaking the trial, including summary of relevant studies (published and unpublished) examining benefits and harms for each intervention | __7-8__ |
|  | 6b | Explanation for choice of comparators | __7-8__ |
| Objectives | 7 | Specific objectives or hypotheses | __7-8____ |
| Trial design | 8 | Description of trial design including type of trial (eg, parallel group, crossover, factorial, single group), allocation ratio, and framework (eg, superiority, equivalence, noninferiority, exploratory) | __7-8___ |
| Methods: Participants, interventions, and outcomes | | |  |
| Study setting | 9 | Description of study settings (eg, community clinic, academic hospital) and list of countries where data will be collected. Reference to where list of study sites can be obtained | __6, 10,*___ |
| Eligibility criteria | 10 | Inclusion and exclusion criteria for participants. If applicable, eligibility criteria for study centres and individuals who will perform the interventions (eg, surgeons, psychotherapists) | __Table 2___ |
| Interventions | 11a | Interventions for each group with sufficient detail to allow replication, including how and when they will be administered | __11-12___ |
|  | 11b | Criteria for discontinuing or modifying allocated interventions for a given trial participant (eg, drug dose change in response to harms, participant request, or improving/worsening disease) | __11-12_ _ |
|  | 11c | Strategies to improve adherence to intervention protocols, and any procedures for monitoring adherence (eg, drug tablet return, laboratory tests) | __11_ _ |
|  | 11d | Relevant concomitant care and interventions that are permitted or prohibited during the trial | __12 _ |
| Outcomes | 12 | Primary, secondary, and other outcomes, including the specific measurement variable (eg, systolic blood pressure), analysis metric (eg, change from baseline, final value, time to event), method of aggregation (eg, median, proportion), and time point for each outcome. Explanation of the clinical relevance of chosen efficacy and harm outcomes is strongly recommended | __15, Table 3 _ |
| Participant timeline | 13 | Time schedule of enrollment, interventions (including any run-ins and washouts), assessments, and visits for participants. A schematic diagram is highly recommended (see Figure) | __Table 1 _ |
| Sample size | 14 | Estimated number of participants needed to achieve study objectives and how it was determined, including clinical and statistical assumptions supporting any sample size calculations | __ 15-16 _ |
| Recruitment | 15 | Strategies for achieving adequate participant enrollment to reach target sample size | ____NA__ _ |
| **Methods: Assignment of interventions (for controlled trials)** | | |  |
| Allocation: |  |  |  |
| Sequence generation | 16a | Method of generating the allocation sequence (eg, computer-generated random numbers), and list of any factors for stratification. To reduce predictability of a random sequence, details of any planned restriction (eg, blocking) should be provided in a separate document that is unavailable to those who enrol participants or assign interventions | __ 10-11_ |
| Allocation concealment mechanism | 16b | Mechanism of implementing the allocation sequence (eg, central telephone; sequentially numbered, opaque, sealed envelopes), describing any steps to conceal the sequence until interventions are assigned | __ 10-11 _ |
| Implementation | 16c | Who will generate the allocation sequence, who will enrol participants, and who will assign participants to interventions | __ 10-11 _ |
| Blinding (masking) | 17a | Who will be blinded after assignment to interventions (eg, trial participants, care providers, outcome assessors, data analysts), and how | __ 10-11 _ |
|  | 17b | If blinded, circumstances under which unblinding is permissible, and procedure for revealing a participant’s allocated intervention during the trial | __ NA_ |
| **Methods: Data collection, management, and analysis** | | |  |
| Data collection methods | 18a | Plans for assessment and collection of outcome, baseline, and other trial data, including any related processes to promote data quality (eg, duplicate measurements, training of assessors) and a description of study instruments (eg, questionnaires, laboratory tests) along with their reliability and validity, if known. Reference to where data collection forms can be found, if not in the protocol | __ 13, supplement_ |
|  | 18b | Plans to promote participant retention and complete follow-up, including list of any outcome data to be collected for participants who discontinue or deviate from intervention protocols | __ 13 |
| Data management | 19 | Plans for data entry, coding, security, and storage, including any related processes to promote data quality (eg, double data entry; range checks for data values). Reference to where details of data management procedures can be found, if not in the protocol | __ 23, supplement_ |
| Statistical methods | 20a | Statistical methods for analysing primary and secondary outcomes. Reference to where other details of the statistical analysis plan can be found, if not in the protocol | __ 17-20 _ |
|  | 20b | Methods for any additional analyses (eg, subgroup and adjusted analyses) | __17-20 _ |
|  | 20c | Definition of analysis population relating to protocol non-adherence (eg, as randomised analysis), and any statistical methods to handle missing data (eg, multiple imputation) | __ 20 _ |
| **Methods: Monitoring** | | |  |
| Data monitoring | 21a | Composition of data monitoring committee (DMC); summary of its role and reporting structure; statement of whether it is independent from the sponsor and competing interests; and reference to where further details about its charter can be found, if not in the protocol. Alternatively, an explanation of why a DMC is not needed | __ 16, supplement |
|  | 21b | Description of any interim analyses and stopping guidelines, including who will have access to these interim results and make the final decision to terminate the trial | __ 16 _ |
| Harms | 22 | Plans for collecting, assessing, reporting, and managing solicited and spontaneously reported adverse events and other unintended effects of trial interventions or trial conduct | supplement _ |
| Auditing | 23 | Frequency and procedures for auditing trial conduct, if any, and whether the process will be independent from investigators and the sponsor | supplement _ |
| Ethics and dissemination | | |  |
| Research ethics approval | 24 | Plans for seeking research ethics committee/institutional review board (REC/IRB) approval | __ 10_ |
| Protocol amendments | 25 | Plans for communicating important protocol modifications (eg, changes to eligibility criteria, outcomes, analyses) to relevant parties (eg, investigators, REC/IRBs, trial participants, trial registries, journals, regulators) | __ 23 _ |
| Consent or assent | 26a | Who will obtain informed consent or assent from potential trial participants or authorised surrogates, and how (see Item 32) | __ NA _ |
|  | 26b | Additional consent provisions for collection and use of participant data and biological specimens in ancillary studies, if applicable | __ NA _ |
| Confidentiality | 27 | How personal information about potential and enrolled participants will be collected, shared, and maintained in order to protect confidentiality before, during, and after the trial | __ supplement_ |
| Declaration of interests | 28 | Financial and other competing interests for principal investigators for the overall trial and each study site | __ supplement_ |
| Access to data | 29 | Statement of who will have access to the final trial dataset, and disclosure of contractual agreements that limit such access for investigators | __ supplement_ |
| Ancillary and post-trial care | 30 | Provisions, if any, for ancillary and post-trial care, and for compensation to those who suffer harm from trial participation | __ NA _ |
| Dissemination policy | 31a | Plans for investigators and sponsor to communicate trial results to participants, healthcare professionals, the public, and other relevant groups (eg, via publication, reporting in results databases, or other data sharing arrangements), including any publication restrictions | __ 23 _ |
|  | 31b | Authorship eligibility guidelines and any intended use of professional writers | __ NA _ |
|  | 31c | Plans, if any, for granting public access to the full protocol, participant-level dataset, and statistical code | __ supplement_ |
| Appendices |  |  |  |
| Informed consent materials | 32 | Model consent form and other related documentation given to participants and authorised surrogates | __ NA _ |
| Biological specimens | 33 | Plans for collection, laboratory evaluation, and storage of biological specimens for genetic or molecular analysis in the current trial and for future use in ancillary studies, if applicable | __ NA _ |

*It is strongly recommended that this checklist be read in conjunction with the SPIRIT 2013 Explanation & Elaboration for important clarification on the items. Amendments to the protocol should be tracked and dated. The SPIRIT checklist is copyrighted by the SPIRIT Group under the Creative Commons “[Attribution-NonCommercial-NoDerivs 3.0 Unported](http://www.creativecommons.org/licenses/by-nc-nd/3.0/)” license.

**2. List of PREOXI Investigators**

Wake Forest School of Medicine- Kevin W. Gibbs, MD*, Jessica A. Palakshappa MD MS*, John P. Gaillard, MD*, Jordan Kugler Goranson, MD*, Madeline Hicks, BS, Haileigh Henson, RN, Savanna Burgess, RN, Benjamin Richards RN, Matthew Strong, RN, Charles Yarbrough, RN, Paul Finkelstein, RN, Tina Lovings, MHS RRT-ACCS, Nikki Barrett, MBA-HM BSRT RRT, Francis Almonor, BS RRT.

Hennepin County Medical Center- Matthew E. Prekker, MD MPH*, Brian E. Driver, MD*, Sydney J. Hansen, MD, Sarah Knack, MD, Leyla Taghizadeh, BA, Kowsar Hurreh, BA, Stephen Douglas, BA, Joshua L. Blaise, RRT, Audrey Hendrickson, MPH.

Vanderbilt University Medical Center- Jonathan D. Casey, MD MSc*, Liza M. Frawley, BSN RN, Margaret A. Hays, RN MSN, Brant Imhoff, MS*, Christina Kampe MA CCRP, Bradley D. Lloyd, RRT RRT-ACCS*, Christopher J. Lindsell, PhD, Li Wang, MS, Tanya K. Marvi, MD, Edward T. Qian, MD, Colleen M. Ratcliff, BS, Jillian P. Rhoads, PhD*, Todd W. Rice, MD, MSc*, Kevin P. Seitz, MD MSc*, Wesley H. Self, MD MPH*, Matthew W. Semler, MD MSc*, Kelsey Womack, PhD*, Jacob A. Wood, BS.

Lahey Hospital & Medical Center- Susan B. Stempek ,MBA MMSc PA*, Joanne M. Wozniak, MS PA*, Andrew Molgaard ,MS PA, AnnMarie Kulis, MHAc RRT, Anthony Campagna, MD, Anthony W. Gray Jr., MD, Avignat Patel, MD, Carla Lamb, MD, Christopher Adler, PA, Christopher Hansen, MD, Conor Crowley, MSN NP, Cynthia Salerno, RN, Daniel Fitelson, MD, Daniel Gavin, RRT, Eleanor Summerhill, MD, Emily Adams, PA, Ezra Miller, MD, Fraser Mackay, MD, Gena Han, DO, Hana Rajevac, MD, Jaclyn Gladstein, PA, Jared Beaudin, DO, Jeffrey Klenz, MD, Joseph Plourde, MPAS, PA, Karl Anderson, DO, Kayla Smith, PA, Kelly Duran, PA, Kevin Clark, MD, Larissa Tian, PA, Laura Mahoney, PA, Luan Nghiem, MD, Mackenzie Stutzman, NP, Marcelous Johnson, MHA RRT-NPS, Michael Colancecco, DO, Michael Plourde, MS PA, Paul Pikman, DO, Rhea Votipka, MSN NP, Robert Spetrini III, DO, Sara Shadchehr, MS DO, Susan Wilcox, MD, Tamar Geva, MD, Victor Pinto-Plata, MD, Vincent Kang, DO.

Ohio State University- Caleb Taylor, MD, MPH*, Matthew C. Exline, MD, MPH*, Sarah Karow, Gabrielle Swoope.

Baylor Scott & White Medical Center- Heath White, DO MS*, Shekhar Ghamande, MD*, Alfredo Vazquez, DO, Alejandro C. Arroliga, MD, Carl Bothel, MD, Tasnim Lat, DO, Hanish Patel, MD, Abirami Subramanian, MD, Calli Bertschy, DO, Eugene Quaye, DO, Adam Letvin, MD, Jennifer Perry, DO, Hannah Shine, DO, Shreyas Boddu, DO, Nithin Kessireddy, MD, Amanpreet Kaur, DO, Zenia Sattar, MD.

University of Colorado School of Medicine- Jason C. Brainard, MD*, Jill Bastman, BSN*,Daniel Resnick-Ault, MD*,Cori Withers, BS*, Neil R. Aggarwal, MD*, Peter D. Sottile, MD*, Erin Anderson, RN*, Kristin Maestas*, Adit A Ginde, MD MPH*, Nicholas J. Kelly, MD, Jenna S. Perrodin, MSN, Danielle Refvem, BSN, Carrie B. Higgins, BSN, Vikhyat Bebarta, MD.

University of Alabama at Birmingham Medical Center and Heersink School of Medicine- Sheetal Gandotra, MD*, Derek W. Russell, MD*, Micah R. Whitson, MD*, David B. Page, MD MSPH*, Anna Altz-Stamm ,RN BSN CCRN, Mary Clay Boone, RN BSN, Robert B. Jonson, RRT, Geri-Anne Warman, RN, BSN, Jennifer J. Oswald, RN, BSN, Jerrod Isbell, RRT, Anne Merril Mason, RN, BSN, Gina White, RN, BSN, Drew Robinson, MD, Jordan Minish, MD, Reed Lahaye, MD, Edwin Gunn MD, Abdulhakim Tlimat, MD, Tyler Greathouse, DO, Luis L. Tatem, MD, Christopher Richardson, MD, Austin Oslock, MD, John Patrick Simmons, MD, Morgan Locy, MD, PhD, Ryan Goetz, MD, Daniel Sullivan, MD, Ross, Schumacher, MD, Melissa Jordan, MD, Jonathan Kalehoff, MD, Anneka Hutton, MD, Daniel Kelmenson, MD, Meena Sridhar, MD, Ahmed Salem, MD, Aneesah B. Jaumally, MD, Ishan Lalani MD, MPH, William S. Stigler MD, Phillip J. O’Reilly, MD, Donna S. Harris, RN, BSN, Cara E. Porter, RN, AND, Sonya Hardy, MA, Puneet Aulakh, MD, Joseph B. Barney, MD, Joseph Chiles III, MD, Bryan Garcia, MD, Aditya Kotecha, MD, Takudzwa Mkorombindo, MD, Peter Morris, MD, Kinner Patel, MD, R. Chad Wade, MD, Carla Copeland, MD, Drew Vestri, RN, BSN, Kelsey Jones, RN, BSN, Regina Oliver, RN, BSN, CCRN, Megan Shelton, MSN, RN, Reagan Isbell, RN, BSN, Lisa Sarratt, Rn, BSN, Sarah W. Robison, MD, Nicole Walker, MS, RRT, Stephanie Powell, RRT, Great Peagler-Mims, MS, RRT, Megan Crumptom, MS, RRT, Laquata Boswell, MS, RRT, Dianne Freeman, BS, RRT.

Oregon Health & Science University - Akram Khan, MD*, Kinsley A. Hubel, MD*,Andrew Connor, MD*, Jesus Martinez, Minn Oh, Genesis Briceno, Jose Pena, Edvinas Pocius, Cassandra Ahmed, Randall Clark.

Montefiore Medical Center- Daniel G. Fein, MD*, Luke Andrea, MD*, Amira Mohamed, MD*, Ari Moskowitz, MD MPH*, Veronika Blinder, DO*, Rishi Malhotra, MD*, Sachin Batra, MD, Melanie Betchen, MD, Christian Lopez-Padilla, MD, Rithvik Balakrishnan, MD, Amy Suhotliv, MD, , Lewis Eisen, MD, Marjan Islam, MD, Samuel Rednor, MD,Ariel L Shiloh, MD,Sabah Boujid, Daniel Ceusters, Gerardo Eman, MD, Ofelia Garcia, Renata Lerner MD, Brenda Y Lopez, Alberto Pinsino, MD, Martha Torres, Aron Solieman, MD, Maykl Mosheyev, MD, Meagan Murphy MD, Ali A. Naqvi MD, Ibrahim Migdady MD.

University of Wisconsin School of Medicine and Public Health- Stephen J. Halliday, MD, MSc*, Micah T. Long MD*, Majid Afshar MD, Amik Sodhi MD.

Brooke Army Medical Center- Brit J. Long, MD*, Steven G. Schauer, DO MS*.

University Medical Center New Orleans and Lousiana State University School of Medicine- David R. Janz MD MSc*.

Denver Health Medical Center-Stacy A. Trent MD MSPH*.

*Denotes an author listed on the byline

**3. Definition of ICU-Free Days (ICU-FDs)**

ICU-FDs are defined as the number of calendar days, between enrollment and 28 days after enrollment, on which the patient is alive and not admitted to an intensive care unit service after the patient’s final discharge from the intensive care unit. Patients who are never

discharged from the intensive care unit receive a value of 0. Patients who die before day 28 receive a value of 0. For patients who are discharged from the ICU, return to an ICU, and are subsequently discharged prior to day 28, ICU-free days are counted from the date of final ICU discharge. All data are censored at hospital discharge or 28 days, whichever comes first.

**4. Definition of Ventilator-Free Days (VFDs)**

VFDs are defined as the number of calendar days, between enrollment and 28 days after enrollment, during which the patient is alive and with unassisted breathing and remains free of assisted breathing. If a patient is liberated from assisted breathing, returns to assisted breathing and subsequently achieves unassisted breathing prior to day 28, VFD will be counted from the end of the last period of assisted breathing to day 28. If the patient is receiving assisted ventilation at day 28 or dies prior to day 28, VFDs are 0. If a patient is discharged while receiving assisted ventilation, VFDs are 0. All data is censored at hospital discharge or 28 days, whichever comes first.

**5. Safety Monitoring and Adverse Events**

Assuring patient safety is an essential component of this protocol. Preoxygenation with non-invasive positive pressure ventilation and preoxygenation with facemask oxygen are both standard-of-care interventions that have been used in clinical practice for decades with an established safety profile. However, any trial conducted during a high-risk, time-sensitive procedure like tracheal intubation of critically ill patients raises unique safety considerations. This protocol addresses these considerations through:

1. Exclusion criteria designed to prevent enrollment of patients likely to experience adverse events from preoxygenation with non-invasive positive pressure ventilation or preoxygenation with facemask oxygen;

2. Systematic collection of outcomes relevant to the safety of preoxygenation with non-invasive positive pressure ventilation and preoxygenation with facemask oxygen;

3. Structured monitoring, assessment, recording, and reporting of adverse events

**5.1. Adverse Event Definitions**

*Adverse Event –* An adverse event will be defined as any untoward or unfavorable medical occurrence in a human subject temporally associated with the subject’s participation in the research, whether or not considered related to the subject’s participation in the research. Any adverse event occurring during the research will be classified according to the following characteristics:

- *Seriousness –* An adverse event will be considered “serious” if it:
  - Results in death;
  - Is life-threatening (defined as placing the patient at immediate risk of death);
  - Results in inpatient hospitalization or prolongation of existing hospitalization;
  - Results in a persistent or significant disability or incapacity;
  - Results in a congenital anomaly or birth defect; or
  - Based upon appropriate medical judgment, may jeopardize the patient’s health and may require medical or surgical intervention to prevent one of the other outcomes listed in this definition.
- *Unexpectedness* – An adverse event will be considered “unexpected” if the nature, severity, or frequency is neither consistent with:
  - The known or foreseeable risk of adverse events associated with the procedures involved in the research that are described in the protocol-related documents, such as the IRB-approved research protocol; nor
  - The expected natural progression of any underlying disease, disorder, or condition of the subject experiencing the adverse event and the subject’s predisposing risk factor profile for the adverse event.
- *Relatedness* – The strength of the relationship of an adverse event to a study intervention or study procedure will be defined as follows:
  - Definitely Related: The adverse event follows (1) a reasonable, temporal sequence from a study procedure AND (2) cannot be explained by the known characteristics of the patient’s clinical state or other therapies AND (3) evaluation of the patient’s clinical state indicates to the investigator that the experience is definitely related to study procedures.
  - Probably or Possibly Related: The adverse event meets some but not all of the above criteria for “Definitely Related”.
  - Probably Not Related: The adverse event occurred while the patient was on the study but can reasonably be explained by the known characteristics of the patient’s clinical state or other therapies.
  - Definitely Not Related: The adverse event is definitely produced by the patient’s clinical state or by other modes of therapy administered to the patient.
  - Uncertain Relationship: The adverse event does not fit in any of the above categories.

**5.2 Monitoring for Adverse Events**

The time interval during which patients will be monitored for the occurrence of adverse events begins at randomization and ends at the first of hospital discharge or 28 days. Adverse events occurring before randomization or after hospital discharge or 28 days will not be collected. The lead investigator at each enrolling site will have primary responsibility overseeing the monitoring, assessment, and reporting of adverse events. Site study personnel will evaluate for the occurrence of adverse events by manual review of the electronic health record and by communication with treating clinicians. Site study personnel will evaluate for the occurrence of adverse events by manual review of the electronic health record at two time points. The first will occur approximately 24 hours after randomization at the time of initial data collection. The second will occur at the first of hospital discharge or 28 days after enrollment at the time of final data collection. Study personnel at each site will also communicate regularly with the treating clinicians who perform tracheal intubation in the study environments between enrollment and 28 days after enrollment to solicit information about any potential adverse events. If study personnel at a site identify a potential adverse event, the lead investigator at the site will be immediately notified. The lead investigator at the site will assess the seriousness, unexpectedness, and relatedness of the potential adverse event. With assistance as needed from the coordinating center and the trial primary investigator, the lead investigator at the site will determine whether the event qualifies for recording and reporting.

**5.3 Recording and Reporting Adverse Events**

The following types of adverse events will be recorded and reported:

- Adverse events that are Serious and Definitely Related, Probably or Possibly Related, or of Uncertain Relationship.
- Adverse events that are Unexpected and Definitely Related, Probably or Possibly Related, or of Uncertain Relationship.

Adverse events that do not meet the above criteria will not be recorded or reported. Adverse events that the lead investigator at a site assesses to meet the above criteria for recording and reporting will be entered into the adverse event electronic case report form in the trial database. The lead investigator at the site will record an assessment of each characteristic for the adverse event, including seriousness, unexpectedness, and relatedness. For any adverse event that is **serious AND unexpected**, and definitely related, probably or possibly related, or of uncertain relationship, the lead investigator at the site will report the adverse event to the coordinating center and the trial primary investigator **within 24 hours** of becoming aware of the adverse event. For any other adverse event requiring recording and reporting, the lead investigator at the study site will report the adverse event to the coordinating center and the trial principal investigator **within 72 hours** of becoming aware of the adverse event. The coordinating center and the trial principal investigator will coordinate with the lead investigator at the site to obtain information about the adverse event regarding each characteristic for the adverse event, including seriousness, expectedness, and relatedness. The lead investigator at the site will be responsible for making final determinations regarding seriousness and unexpectedness. The coordinating center and trial principal investigator will be responsible for making final determinations regarding relatedness.

For adverse events that meet the above criteria for recording and reporting, the coordinating center will notify the DSMB, the sIRB, and the sponsor in accordance with the following reporting plan:

| **Characteristics of the Adverse Event** | **Reporting Period** |
| --- | --- |
| Fatal or life-threatening (and therefore serious), unexpected, and definitely related, probably or possibility related, or of uncertain relationship. | Report to the DSMB, sIRB, and sponsor within 7 days after notification of the event. |
| Serious but non-fatal and non-life-threatening, unexpected, and definitely related, probably or possibly related, or of uncertain relationship. | Report to DSMB, sIRB, and sponsor within 15 days of notification of the event. |
| All other adverse events meeting criteria for recording and reporting. | Report to DSMB in regularly scheduled DSMB safety reports. |

**5.4 Clinical Outcomes that may be Exempt from Adverse Event Recording and Reporting**

In this study of critically ill patients at high risk for death and other adverse outcomes due to their underlying critical illness, clinical outcomes, including death and organ dysfunction, will be systematically collected and analyzed for all patients. The primary, secondary, safety, and exploratory outcomes will be recorded and reported as clinical outcomes and not as adverse events unless treating clinicians or site investigators believe the event is Definitely Related or Probably or Possibly Related to the study intervention or study procedures. This approach – considering death and organ dysfunction as clinical outcomes rather than adverse events and systemically collecting these clinical outcomes for analysis – is common in ICU trials. This approach ensures comprehensive data on death and organ dysfunction for all patients, rather than relying on sporadic adverse event reporting to identify these important events. The following events are examples of study-specific clinical outcomes that would not be recorded and reported as adverse events unless treating clinicians or site investigators believe the event was Definitely Related or Probably or Possibly Related to the study intervention or study procedures:

- Death (all deaths occurring prior to hospital discharge or 28 days will be recorded);
- Organ dysfunction
  - Pulmonary – hypoxemia, aspiration, acute hypoxemic respiratory failure, pneumothorax
  - Cardiac – hypotension, shock, vasopressor receipt, cardiac arrest;
- Duration of mechanical ventilation;
- Duration of ICU admission;
- Duration of hospitalization

Note: A study-specific clinical outcome may also qualify as an adverse event meeting criteria for recording and reporting. For example, a pneumothorax that the investigator considers Definitely Related to preoxygenation with non-invasive positive pressure ventilation would be both recorded as a study-specific clinical outcome and recorded and reported as a Serious and Definitely Related adverse event.

**5.5 Unanticipated Problems involving Risks to Subjects or Others**

Investigators must also report Unanticipated Problems Involving Risks to Subjects or Others (“Unanticipated Problems”), regardless of severity, associated with study procedures **within 24 hours** of the site investigator becoming aware of the Unanticipated Problem. An Unanticipated Problem is defined as any incident, experience, or outcome that meets all of the following criteria:

- Unexpected (in terms of nature, severity, or frequency) given (a) the research procedures that are described in the protocol-related documents, such as the IRB-approved research protocol; and (b) the characteristics of the subject population being studied; AND
- Definitely Related or Probably or Possibly Related to participation in the research (as defined above in the section on characteristics of adverse events); AND
- Suggests that the research places subjects or others at a greater risk of harm (including physical, psychological, economic, or social harm) than was previously known or recognized.

If any study personnel at a site become aware of an event that may represent an Unanticipated problem, they will immediately contact the lead investigator for the site. The lead investigator for the site will assess whether the event represents an Unanticipated Problem by applying the criteria described above. If the lead investigator at a site determines that the event represents an Unanticipated Problem, the lead investigator at the site will record the Unanticipated Problem in the Unanticipated Problem electronic case report form in the trial database. The lead investigator at the site will then communicate that an Unanticipated Problem has occurred to the coordinating center and the principal investigator **within 24 hours** of the lead investigator at the site becoming aware of the Unanticipated Problem. The coordinating center and principal investigator will coordinate with the lead investigator at the site to obtain information about the Unanticipated Problem. The coordinating center will report the Unanticipated Problem to the DSMB, sIRB, and sponsor within 15 days of becoming aware of the Unanticipated Problem.

**6. Patient Privacy and Data Storage**

At no time during this study, its analysis, or its publication, will patient identities be revealed in any manner. The minimum necessary data containing patient or provider identities or other private healthcare information (PHI), is collected. All subjects are assigned a unique study ID number for tracking purposes. Data collected from the medical record is entered into the secure online database REDCap. The PHI required to accurately collect clinical and outcomes data is available only to investigators at the site at which the subject is enrolled, and this data is shared only in completely de-identified form with the coordinating center via the secure online database REDCap. Hard copies of the data collection sheet completed at the time of the airway management event are stored in a locked room until after the completion of enrollment and data cleaning. Once data are verified and the database is locked, all hard copies of data collection forms will be destroyed. The de-identified dataset housed in REDCap will be accessed by the coordinating center for reporting the results of this trial. All data will be maintained in the secure online database REDCap until the time of study publication. At the time of publication, all PHI at local centers will be expunged and only the de-identified version of the database will be retained. Potential future use of de-identified data generated in the course of this study by the coordinating center and other participating sites is allowed and will be governed by mutual data sharing use agreements. **7. Plan for Communication of Protocol Changes**

Any changes to the trial protocol including changes to eligibility, outcomes and analyses will be implemented via a new version of the full trial protocol, tracked with the date of the update, and the version number of the trial protocol. A list summarizing the changes made with each protocol revision will be included at the end of each protocol. The updated protocol will be sent to the sIRB for approval and tracking prior to implementation of the protocol change. At the time of publication, the original trial protocol, and the final trial protocol, including the summary of changes made with each protocol change, will be included in the supplementary material for publication.

**8. Training Materials**

Prior to initiation of enrolment at participating sites, research personnel and clinicians are trained in best practices regarding preoxygenation with noninvasive ventilation and preoxygenation with an oxygen mask via a standardized training video, which is accessible at <https://www.pragmaticcriticalcare.org>.

The training included the following information relevant to preoxygenation with an oxygen mask: (1) an overview of the types of oxygen masks and the components of each; (2) a recommendation to use the highest available oxygen flow rate (“flush rate oxygen”; ≥ 15 liters per minute) for ≥ 3 minutes for preoxygenation; (3) performance of a jaw thrust maneuver after induction of anesthesia to ensure airway patency; (4) continued provision of supplemental oxygen between induction of anesthesia and initiation of laryngoscopy; (5) the optional administration of manual ventilation between induction and laryngoscopy with a bag-mask device.

The training included the following information relevant to preoxygenation with noninvasive ventilation: (1) an overview of the use of conventional mechanical ventilators or dedicated noninvasive ventilators to provide noninvasive ventilation during preoxygenation; (2) the modes of ventilation capable of delivering noninvasive mechanical ventilation on a conventional mechanical ventilator; (3) an overview of the terminology for inspiratory pressure and expiratory pressure on each device; (4) a recommendation to use an expiratory pressure of ≥ 5 cm H_2_O and an inspiratory pressure of ≥ 10 cm H_2_O and respiratory rate of 10 breaths per minute; (5) ≥ 3 minutes for preoxygenation; (6) performance of a jaw thrust maneuver after induction of anesthesia to ensure airway patency; (7) continuation of noninvasive ventilation between induction of anesthesia and initiation of laryngoscopy.

**9. Supplementary Table 1**

| Supplement Table 1: Study variables | |
| --- | --- |
| **Baseline** | Age |
|  | Sex |
|  | Race and Ethnicity |
|  | Height, weight, and body mass index (BMI) |
|  | Acute Physiology and Chronic Health Evaluation (APACHE II) score^1^ |
|  | Active medical problems at the time of enrollment |
|  | Comorbidities |
|  | Indication for intubation |
|  | Vasopressor receipt in the hour prior to enrollment |
|  | Highest FiO_2_ in the hour prior to enrollment |
|  | Lowest SpO_2_/FiO_2_ ratio (or PaO_2_/ FiO_2_ ratio) in the hour prior to enrollment |
|  | Glasgow Coma Scale score |
|  | Oxygen delivery device at enrollment |
|  | Presence of difficult airway characteristics (limited mouth opening, small mandible, large tongue, short neck, large neck circumference, limited anatomic neck mobility, cervical immobilization due to trauma, obesity) |
|  | Operator’s level of training and specialty |
|  | Operator’s prior intubation experience |
| **Peri-procedural** | SpO_2_ and FiO_2_ at enrollment |
|  | Lowest SpO_2_ from enrollment to induction |
|  | Approach to preoxygenation |
|  | Duration of preoxygenation |
|  | Noninvasive inspiratory and expiratory pressures during preoxygenation |
|  | Time of sedative administration (induction) |
|  | Sedative agent and dose |
|  | Neuromuscular blocking agent and dose |
|  | Administration of a vasopressor prior to induction |
|  | SpO_2_ at induction |
|  | Systolic blood pressure at induction |
|  | Approach to oxygen administration and ventilation between induction and laryngoscopy |
|  | Time of start of first laryngoscopy attempt |
|  | Laryngoscope model, blade size, blade shape on first attempt |
|  | Use of video screen (if applicable) on first attempt |
|  | Best Cormack-Lehane grade of glottis view on first laryngoscopy attempt |
|  | Presence of body fluids obstructing laryngeal view |
|  | Presence of upper airway obstruction or edema |
|  | Receipt of chest compressions at time of first laryngoscopy attempt |
|  | Number of intubation attempts (number of times laryngoscope entered mouth) |
|  | Number of times bougie entered mouth (if applicable) |
|  | Number of times endotracheal tube entered mouth |
|  | Reason for failure of first intubation attempt (if applicable) |
|  | Procedural adjustments made for the final intubation attempt (if applicable) |
|  | Necessity of an additional operator |
|  | Esophageal intubation |
|  | Injury to the teeth |
|  | Operator-reported aspiration between induction and intubation |
|  | Time of successful tracheal intubation |
|  | Endotracheal tube size |
|  | Lowest SpO_2_ from induction until 2 minutes after intubation |
|  | Cardiac arrest from induction until 2 minutes after intubation not resulting in death within 1 hour of induction |
|  | Cardiac arrest from induction until 2 minutes after intubation resulting in death within 1 hour of induction |
| **In-hospital** |  |
| *24 hours after enrollment* | New pneumothorax detected in first 24 hours after enrollment |
|  | New infiltrate on chest imaging in the 24 hours after intubation |
|  | Vasopressor receipt at 24 hours after enrollment |
|  | SpO_2_ at 24 hours after enrollment |
|  | FiO_2_ at 24 hours after enrollment |
|  | PEEP at 24 hours after enrollment |
|  | Systolic blood pressure at 24 hours after enrollment |
|  | Number of attempts at passing a bougie. |
|  | Number of attempts at passing an endotracheal tube. |
|  | Incidence of cardiovascular collapse, defined as a composite of one or more of the following between induction and 2 minutes after intubation: Systolic blood pressure <65 mmHg, new or increased vasopressor, cardiac arrest not resulting in death within 1 hour of induction, cardiac arrest resulting in death within 1 hour of induction. |
| *28 days after enrollment* | 28 day in-hospital mortality. |
|  | Ventilator-free days to 28 days. |
|  | ICU-free days to 28 days. |

1.	Knaus WA, Draper EA, Wagner DP, Zimmerman JE. APACHE II: a severity of disease classification system. *Crit Care Med*. Oct 1985;13(10):818-29.
